# Urban versus rural prevalence of intestinal parasites using multi-parallel qPCR in Colombia

**DOI:** 10.1101/2020.09.20.20197616

**Authors:** Patricia E. Bryan, Marcela Romero, Miryan Sánchez, Giovanny Torres, Wilber Gómez, Marcos Restrepo, Alejandro Restrepo, Rojelio Mejia

**Affiliations:** Laboratory of Human Parasitology, National School of Tropical Medicine, Baylor College of Medicine, Houston, Texas; Instituto Colombiano de Medicina Tropical, Universidad CES, Medellín, Colombia

## Abstract

Stool samples from 122 children from urban slum (n = 72) and rural (n = 50) areas were analyzed using multi-parallel real-time qPCR to determine the prevalence of intestinal parasites from two communities in Colombia. Findings indicated a prevalence of 86.1% *Blastocystis* spp., 62.5% *Giardia intestinalis*, 19.4% *Cryptosporidium* spp., 19.4% *Ascaris lumbricoides*, and 5.6% *Trichuris trichiura* in an urban slum; and 76% *Blastocystis* spp., 68% *Giardia intestinalis*, 20% *Entamoeba histolytica*, 50% *Ascaris lumbricoides*, 46% *Trichuris trichiura* and 2% *Strongyloides stercoralis* in rural areas. Polyparasitism was higher in rural (58%) compared to urban (25%) areas (*p* = 0.001). *Trichuris trichiura* burden was higher in the rural area (*p* = 0.002). Over 40% of helminth infections in rural areas had a heavy parasite burden by WHO classification. Over half of urban and rural children were infected with *Giardia intestinalis* and *Blastocystis* spp. Our data provides accurate epidemiologic surveillance for public health interventions.

---

Intestinal parasites are globally widespread infectious organisms disproportionately affecting children in resource-limited areas with associated morbidity that is poorly understood and can have long-term child health implications. Environmental surroundings influence exposure to these parasites, as does the differences in communities. Reliable, highly sensitive and specific diagnostic tests for intestinal parasitic infections are critical for treatment decisions in mass drug administration programs, impact evaluation, and surveillance.^1-3^

A multi-parallel real-time quantitative PCR (qPCR) assay was used to detect intestinal parasites commonly infecting children living in resource-limited areas including soil-transmitted helminths (STH) (*Ascaris lumbricoides, Ancylostoma duodenale, Necator americanus, Strongyloides stercoralis*, and *Trichuris trichiura*), protozoa (*Cryptosporidium* spp., *Entamoeba histolytica*, and *Giardia intestinalis*) and heterokont (*Blastocystis* spp.). Data from Colombia on intestinal parasite prevalence in children is limited, especially among preschool-age children.^4, 5^ The study population included preschool and school-age children living in urban slums of the city of Medellín (mean age = 2 years) and rural areas of the town of Unguía (mean age = 2.5 years) located in the northwest Andean region of Colombia. Recently, protozoa detected by a molecular approach was reported among school-age children in an urban area of southwest Colombia with a prevalence of 39.2% *Blastocystis* spp., 10.6% *G. intestinalis*, and 9.8% *Cryptosporidium* spp.^3^ Intestinal parasites were previously detected by microscopy among children under 15 years of age in a rural area of the northern coastal region of Colombia with 63% infected with protozoa and 69% infected with STH.^6^ In Colombia, studies reporting molecular epidemiologic data on intestinal parasites among children from both urban and rural settings in the same study population are sparse.^3,7-9^ Limited molecular epidemiologic surveillance data on childhood intestinal parasitism in areas with concentrations of poverty undermines positive child health outcomes.^3,9^ Our study contributes to data comparing the prevalence of intestinal parasites detected by qPCR among urban slum and rural children from the study sampling areas in Colombia. Because the interplay between socioeconomic and environmental factors, hygiene, and transmission dynamics contribute to increased child vulnerability to intestinal parasite exposure.^10^ accurate epidemiologic data on both STH and protozoa is critical to decision making for treatment and public health interventions for populations of children in contrasting community settings.

Stool samples were analyzed from 122 Colombian children living in urban slums (n = 72) and rural areas (n = 50). Parasite DNA was extracted from 50 mg of stool from each stool sample using MP FastDNA™ for Soil Kit (MP Biochemicals, Solon, OH) according to the manufacturer’s instructions for all parasites, except *Trichuris trichiura*, which required an additional heating step.^11^ Species-specific primers and probes for these nine parasites were previously designed and tested demonstrating 100% sensitivity and specificity.^11^ Calculated *G. intestinalis* cysts and STH eggs were derived from spiked samples with known parasite concentrations.^11^

Overall, intestinal parasites among children in this study were found to be similarly highly prevalent in both urban slum (97.2%) and rural areas (90%), with *G. intestinalis* and *Blastocystis* spp. predominantly prevalent across these sampling settings. Because urban populations generally have better access to sanitation and clean water compared to rural populations,^4^ the high prevalence of intestinal parasites; namely *G. intestinalis* and *Blastocystis* spp., in both urban and rural areas (62.5% and 86.1% versus 68% and 76%, respectively) is noteworthy. Previously reported data from Colombia on urban slum children in Medellín indicated a significantly lower prevalence of giardiasis (25.9%) detected by microscopy,^12^ and the most recent national survey reported that 15.4% of Colombian children were infected with *G. intestinalis* and 57.7% were infected with *Blastocystis* spp. also detected by microscopy-based diagnostic methods.^13^ Despite the high prevalence of *Blastocystis* spp., the clinical importance of this parasite remains unclear.^14^ However, this parasite is an important indicator of fecal contamination of food and water.^14^ *G. intestinalis* represents a significant public health problem worldwide.^3, 5^ Giardiasis can cause acute or chronic diarrhea but is often asymptomatic and is associated with detrimental impacts to growth and development in children.^5,14^ In the present study, *G. intestinalis* spiking studies were also conducted, from which cysts per gram of stool were calculated. Subsequently, similar *G. intestinalis* infection burden was found among urban slum and rural children (Figure 1), suggesting exposure to similar risk factors for giardiasis in both settings. Because fecal contamination of drinking water is the most common source of *G. intestinalis* cysts from domestic animal and human origins,^5, 14^ the inadequate or lack of access to clean water typically found in resource-deprived urban slum and rural areas of developing countries may explain the prevalence and parasite burden in the study population.

**Figure 1.**
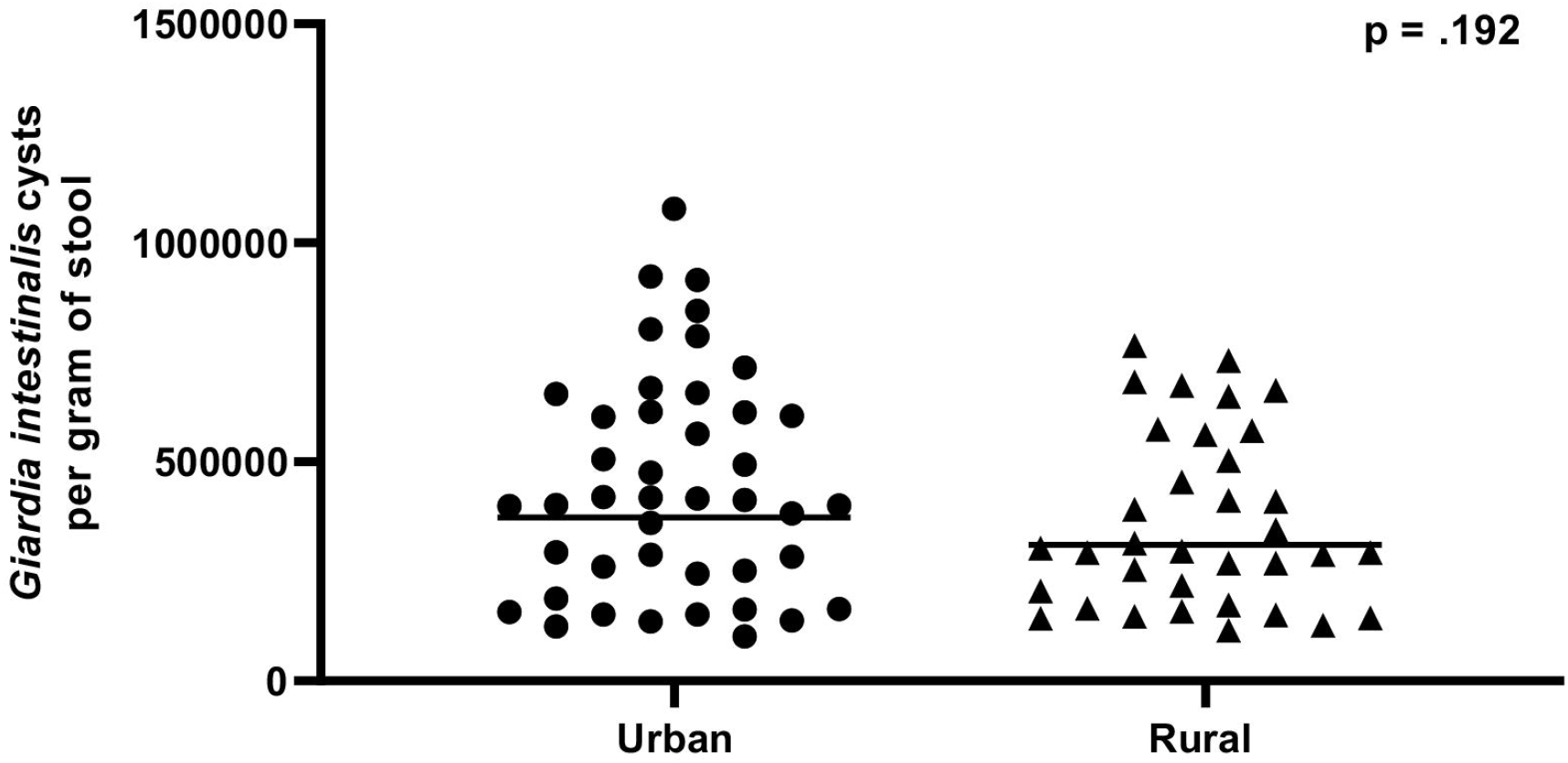
Estimated Giardia intestinalis cysts among children in urban (Medellín) compared to rural area (Unguía) had similar burden of infection.

In this study, helminthiasis was more prevalent among rural children compared to the urban study population. The most prevalent STH, *A. lumbricoides* and *T. trichiura*, were significantly higher among rural children comparably (50% and 46% versus 19.4% and 5.6%, respectively). This was not unexpected due to similar transmission dynamics and greater risk of exposure to contaminated soil in rural settings combined with poor hygiene awareness and hand-to-mouth behavior characteristic of early childhood^10^ To evaluate STH parasite burden, DNA fg/µL was correlated with eggs per gram (epg) of stool-based on WHO threshold criteria for the classification of parasite burden for *A. lumbricoides* (1 – 5,000 light; > 5,000 – 50,000 moderate; and > 50,000 heavy); *T. trichiura* (1 – 999 light; 1,000 – 9,999 moderate; ≥ 10,000 heavy; and for hookworm (1 – 1,999 light; 2,000 – 3,999 moderate; ≥ 4,000 heavy).^15^ Over 40% of STH infections among children in the rural setting were classified as having a heavy parasite burden. Among rural children with trichuriasis, parasite burden was higher (9,953 epg) compared to those in urban slums (325 epg) (p = 0.002) (Figure 2). Polyparasitism was also found in the overall study population, with the highest prevalence found among rural children (58%) compared to children in urban slum areas (25%) (*p* = 0.001). Because morbidity is proportional to parasite burden,^2,9^ polyparasitism in the study population is a significant concern given the young age of these children. This young age group is at increased vulnerability to the detrimental health impacts of intestinal parasites due to critical stages of growth and development occurring during early childhood.^16^ Moreover, polyparasitism involving both STH and protozoa serves as an important indicator of inadequate sanitary conditions and continual reinfection.^9^

**Figure 2.**
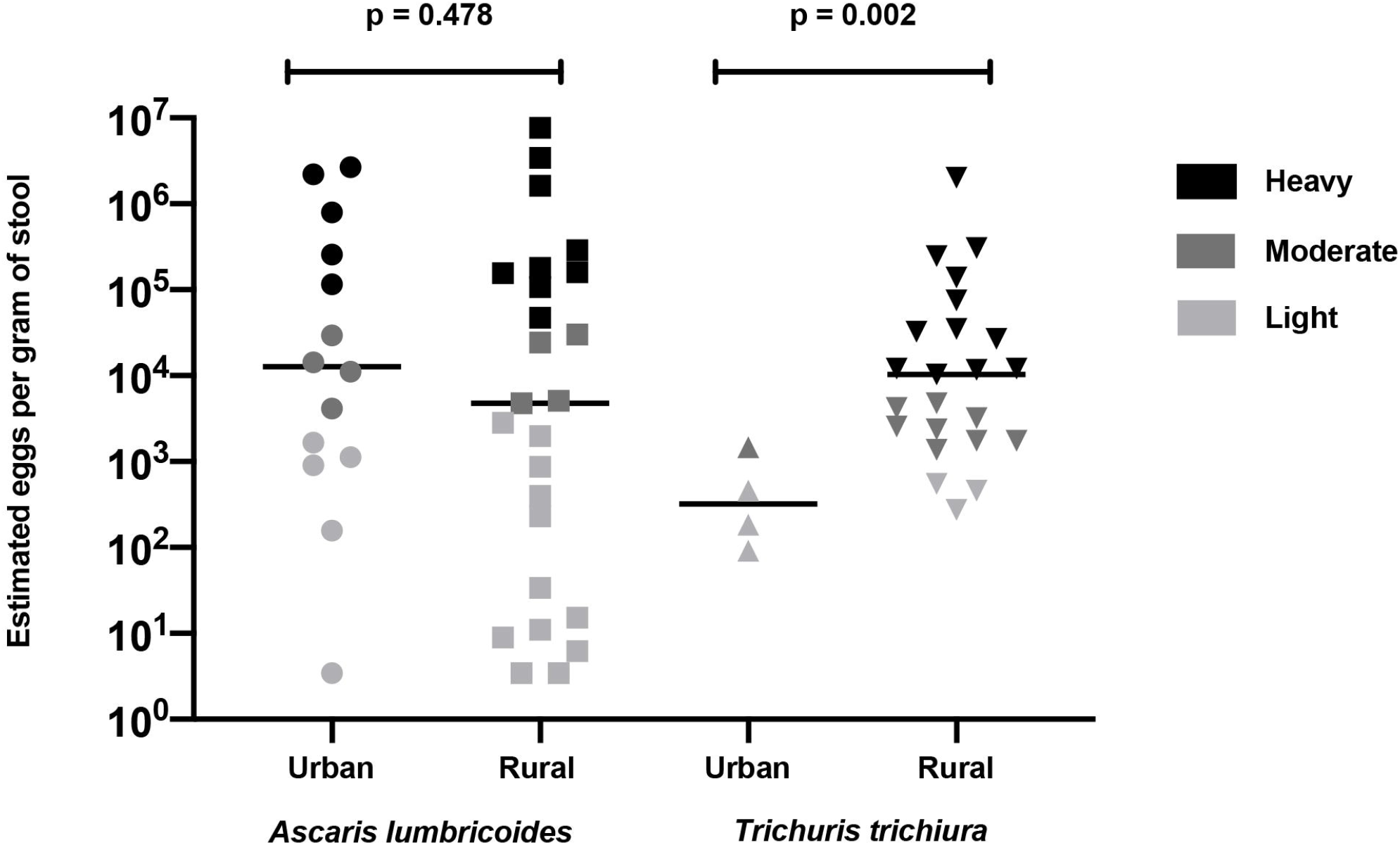
Estimated helminth burden in eggs per gram of stool using qPCR among children in urban (Medellín) compared to rural areas (Unguía) had equal Ascaris but higher Trichuris burden of infection in rural areas.

**Table 1.** Urban (Medellín) and rural (Unguía) parasite prevalence and infection burden in DNA fg/µL.

| Parasite species | Urban<br>(Medellin)<br>n = 72 | Rural<br>(Unguía)<br>n = 50 | *P-value* | Urban DNA<br>(fg/μL)<br>Geomean | Rural DNA<br>(fg/μL)<br>Geomean | *P-value* |
| --- | --- | --- | --- | --- | --- | --- |
| Any parasite | 70 (97.2%) | 45 (90%) | 0.091 |  |  |  |
| *Ascaris lumbricoides* | 14 (19.4%) | 25 (50%) | 0.0004 | 18.20 | 4.764 | 0.478 |
| *Ancylostoma duodenale* | 0 | 0 | * | * | * | * |
| *Necator americanus* | 0 | 0 | * | * | * | * |
| *Strongyloides stercoralis* | 0 | 1 (2%) | * | * | 3.122 | * |
| *Trichuris trichiura* | 4 (5.6%) | 23 (46%) | 0.0001 | 0.0036 | 0.1090 | 0.002 |
| *Blastocystis* spp. | 62 (86.1%) | 38 (76%) | 0.155 | 1.042 | 1.880 | 0.873 |
| *Cryptosporidium* spp. | 14 (19.4%) | 0 | * | 5.576 | * | * |
| *Entamoeba histolytica* | 0 | 10 (20%) | * | * | 2.701 | * |
| *Giardia intestinalis* | 45 (62.5%) | 34 (68%) | 0.533 | 4.720 | 0.7736 | 0.114 |

Findings from this study suggest that children from urban slums of Medellín and rural areas of Unguía in Colombia live in highly contaminated environments with continuous exposure to intestinal parasites in these different communities. The peripheral neighborhoods of the urban poor in cities such as Medellín, Colombia, typically have informal housing development with poor housing standards, overcrowding, and inadequate infrastructure for sanitation and access to clean water.^17-19^ These conditions are comparable to resource-deprived rural settings and can often lead to even greater exposure to fecal contamination in households, schools, and where urban slum children play and spend most of their time.^13^ The predominance of *G. intestinalis* in both urban slum and rural areas suggests the likelihood of exposure to similar contamination sources in geographically contrasting communities.^17^ The high prevalence of intestinal parasites found in the study population confirms that childhood intestinal parasitic infections are a relevant health issue in both urban poor and rural communities in Colombia and serves as an important indicator of community-level socioeconomic development.^8,10^ Because accurate prevalence data in a region or community is essential for identifying local vulnerabilities,^4,17,19^ molecular epidemiologic data from this study provides evidence of important community needs common to both urban slum and rural communities included in this study. Accurate epidemiologic data from this study is also critical for treatment decisions in mass drug administration programs, impact evaluation, and surveillance. This is particularly important given that treatment programs for intestinal parasites in Colombia are not aimed at the preschool-age population and do not include treatment for intestinal protozoa.^4,5^ Moreover, prevalence data generated from this study provides evidence for local decision making for implementing resources to improve living conditions for urban and rural populations of children in Colombia.^14,19^ A major limitation of this study was the lack of *G. intestinalis* assemblages and *Blastocystis* subtype identification. Future studies will include the assemblage and subtyping to allow correlation to human disease.

## Data Availability

All relevant data is included in the paper.

